# Clinical presentation and differential splicing of *SRSF2, U2AF1* and *SF3B1* mutations in patients with Acute Myeloid Leukaemia

**DOI:** 10.1101/2020.01.07.20016881

**Authors:** Stefanos A. Bamopoulos, Aarif M. N. Batcha, Vindi Jurinovic, Maja Rothenberg-Thurley, Hanna Janke, Bianka Ksienzyk, Julia Philippou-Massier, Alexander Graf, Stefan Krebs, Helmut Blum, Stephanie Schneider, Nikola Konstandin, Maria Cristina Sauerland, Dennis Görlich, Wolfgang E. Berdel, Bernhard J. Woermann, Stefan K. Bohlander, Stefan Canzar, Ulrich Mansmann, Wolfgang Hiddemann, Jan Braess, Karsten Spiekermann, Klaus H. Metzeler, Tobias Herold

**Affiliations:** Laboratory for Leukemia Diagnostics, Department of Medicine III, University Hospital, LMU Munich, Munich, Germany; Department of Hematology and Oncology (CBF), Charité University Medicine, Berlin, Germany; Institute for Medical Information Processing, Biometry and Epidemiology, LMU Munich, Munich, Germany; DIFUTURE (Data integration for Future Medicine (DiFuture, www.difuture.de), LMU Munich, Munich, Germany; Laboratory for Functional Genome Analysis (LAFUGA), Gene Center, LMU Munich, Munich, Germany; Institute of Human Genetics, University Hospital, LMU Munich, Munich, Germany; Institute of Biostatistics and Clinical Research, University of Münster, Münster, Germany; Department of Medicine, Hematology and Oncology, University of Münster, Münster, Germany; German Society of Hematology and Oncology, Berlin, Germany; Leukaemia and Blood Cancer Research Unit, Department of Molecular Medicine and Pa- thology, University of Auckland, Auckland, New Zealand; Gene Center, LMU Munich, Munich, Germany; German Cancer Consortium (DKTK), Partner Site Munich, Munich, Germany; German Cancer Research Center (DKFZ), Heidelberg, Germany; Department of Oncology and Hematology, Hospital Barmherzige Brüder, Regensburg, Germany; Research Unit Apoptosis in Hematopoietic Stem Cells, Helmholtz Zentrum München, German Center for Environmental Health (HMGU), Munich, Germany

## Abstract

Previous studies demonstrated that splicing factor mutations are recurrent events in hematopoietic malignancies with both clinical and functional implications. However, their aberrant splicing patterns in acute myeloid leukaemia remain largely unexplored. In this study we characterized mutations in *SRSF2, U2AF1* and *SF3B1*, the most commonly mutated splicing factors. In our clinical analysis of 2678 patients, splicing factor mutations showed inferior relapse-free and overall survival, however, these mutations did not represent independent prognostic markers. RNA-sequencing of 246 and independent validation in 177 patients revealed an isoform expression profile highly characteristic for each individual mutation, with several isoforms showing a strong dysregulation. By establishing a custom differential splice junction usage pipeline we accurately detected aberrant splicing in splicing factor mutated samples. Mutated samples were characterized predominantly by decreased junction usage. A large proportion of differentially used junctions were novel. Targets of splicing dysregulation included several genes with a known role in leukaemia. In *SRSF2*(P95H) mutants we further explored the possibility of a cascading effect through the dysregulation of the splicing pathway. We conclude that splicing factor mutations do not represent independent prognostic markers. However, they do have genome-wide consequences on gene splicing leading to dysregulated isoform expression of several genes.

## Introduction

The discovery of recurring somatic mutations within splicing factor genes in a large spectrum of human malignancies has brought attention to the critical role of splicing and its complex participation in carcinogenesis [1–3]. The spliceosome is a molecular machine assembled from small nuclear RNA (snRNA) and proteins and is responsible for intron removal (splicing) in pre-messenger RNA. In acute myeloid leukaemia (AML), splicing factor mutations occur most frequently in *SRSF2, U2AF1* and *SF3B1*. The splicing factors encoded by these genes are all involved in the recognition of the 3’-splice site during pre-mRNA processing.[4] Splicing factor (SF) mutations are especially common in haematopoietic malignancies, where they occur early on and remain stable throughout the disease evolution of myelodysplastic syndromes (MDS) [1,5–9]. SF mutations are also prevalent in acute myeloid leukaemia (AML), which is often the result of myeloid dysplasia progression, with reported frequencies of 6-10%, 4-8% and 3% for *SRSF2, U2AF1* and *SF3B1* mutations respectively [2,4,10,11].

SF mutations rarely co-occur within the same patient, implying the lack of a synergistic effect or synthetic lethality [1,2,6]. They are typically heterozygous point mutations, frequently coincide with other recurrent mutations in haematopoietic malignancies and are associated with aberrant splicing in genes recurrently mutated in AML [2,4,8]. Notably, the aberrant splicing patterns are distinct for each SF mutation, suggesting that SF mutations do not share the same mechanism of action and should be recognized as individual alterations [4,9,12–17]. The clinical characteristics and outcome of patients with SF mutations are well defined in MDS [1,3,8,9]. Meanwhile, attempts at determining the role of SF mutations as independent prognostic markers in AML have often been limited to specific subgroups and it remains unclear whether the inferior survival associated with SF mutations is confounded by their association with older age or accompanying mutations [10,18]. Additionally, while evidence of aberrant splicing due to SF mutations has emerged for many genes relevant in AML, it is yet uncertain whether and how these changes directly influence disease initiation or evolution.

The aim of this study was a comprehensive analysis of the prognostic implications of SF mutations in two well-characterized and intensively treated adult AML patient cohorts amounting to a total of 2678 patients. In addition, the core functional consequences of SF mutations were explored using targeted amplicon sequencing in conjunction with RNA-sequencing on two large datasets.

## Patients and Methods

### Patients

Our primary cohort included a total of 1138 AML patients treated with intensive chemotherapy in two randomized multicenter phase 3 trials of the German AML Cooperative Group (AMLCG). Treatment regimens and inclusion criteria are described elsewhere [2]. A cohort of 1540 AML patients participating in multicenter clinical trials of the German-Austrian AML Study Group (AMLSG), was used for validation [19]. Cohort composition and filtering criteria are outlined in the supplementary.

### Molecular Workup

All participants of the AMLCG cohort received cytogenetic analysis, as well as targeted DNA-sequencing as described previously [2]. The subset of the AMLSG cohort included in this study received a corresponding molecular workup, described elsewhere [19].

### RNA-Sequencing and data processing

Using the Sense mRNA Seq Library Prep Kit V2 (Lexogen; Vienna, Austria) 246 samples underwent poly(A)-selected, strand-specific, paired-end sequencing on a HiSeq 1500 instrument (Illumina; San Diego, CA, USA). A subset of the Beat AML cohort (n=177) was used for validation [20]. The same bioinformatics analysis was used for both datasets and is described in the supplementary. The samples were aligned to the reference genome (Ensembl GRCh37 release 87) using the STAR [21] aligner with default parameters. Splice junctions from all samples were pooled, filtered and used to create a new genomic index. Multi-sample 2-pass alignments to the re-generated genome index followed, using the STAR recommended parameters for gene-fusion detection. Read counts of transcripts and genes were measured with salmon [22]. Read counts of splice junctions were extracted from the STAR output.

### Differential expression analysis and differential splice junction usage (DSJU)

A minimum expression filter was applied prior to each differential analysis. Differentially expressed isoforms were identified with the limma [23] package after TMM-normalization [24] with edgeR [25] and weighting with voom [26,27]. A surrogate variable analysis step using the sva [28] package was included to reduce unwanted technical noise. DSJU was quantified similarly using the diffSplice function of the limma package. Both analyses are described in detail in the supplementary.

### Nanopore cDNA sequencing and analysis

Total RNA was transcribed into cDNA using the TeloPrime Full-Length cDNA Amplification Kit (Lexogen) which is highly selective for polyadenylated full-length RNA molecules with 5’-cap structures. Two barcoded samples for multiplexed analysis were sequenced on the Oxford Nanopore Technologies MinION platform. Alternative isoform analysis was performed with FLAIR [29].

### Statistics

Statistical analysis was performed using the R-3.4.1 [30] software package. Correlations between variables were performed using the Mann-Whitney U test and the Pearson’s chi-squared test. In case of multiple testing, p-value adjustment was performed as described in the supplementary. Survival analysis was performed and visualized using the Kaplan-Meier method and the log-rank test was utilized to capture differences in relapse free survival (RFS) and overall survival (OS). Patients receiving an allogeneic stem cell transplant were censored at the day of the transplant, for both RFS and OS. Additionally, Cox regressions were performed for all available clinical parameters and recurrent aberrations. Cox multiple regression models were then built separately for RFS and OS, using all variables with an unadjusted p-value < 0.1 in the single Cox regression models.

## Results

### Clinical features of AML patients with SF mutations

We characterized SF mutations in two independent patient cohorts (the AMLCG and AMLSG cohorts). Our primary cohort (AMLCG) consisted of 1119 AML patients (Figure S1), 236 (21.1%) of which presented with SF mutations. The three most commonly affected SF genes, *SRSF2, U2AF1* and *SF3B1* were mutated in 12.1% (n=136), 3.4% (n=38) and 4.1% (n=46) of the patients (Figure 1A). In agreement with previous findings [19], SF mutations were in their majority mutually exclusive, heterozygous hotspot mutations (Figure 1B). The four most common point mutations were *SRSF2*(P95H) (n=69), *SRSF2*(P95L) (n=27), *U2AF1*(S34F) (n=18), and *SF3B1*(K700E) (n=18) mutations (Figure 1C-E). The clinical characteristics of patients harboring SF mutations are summarized in Tables 1 and S1 (AMLSG cohort), along with a statistical assessment between cohorts (Table S2). We observed a high overall degree of similarity regarding clinical features of SF mutated patients between the AMLCG and AMLSG cohorts, despite their large median age difference. Mutations in *SRSF2, U2AF1* and *SF3B1* occurred more frequently in secondary AML (44.7% compared to 18.2% in *de novo* AML) and were all associated with older age. As reported previously [1], *SRSF2* and *U2AF1* mutated patients were predominantly male (76.7% and 76.3%, respectively). Furthermore, patients harboring *SRSF2* mutations presented with a lower white blood cell count (WBC; median 13.3 10^9^/L vs. 22.4 10^9^/L) while *U2AF1* mutated patients presented with a reduced blast percentage in their bone marrow when compared to SF wildtype patients (median 60% vs 80%).

**Table 1:** Clinical characteristics of SF mutations in the AMLCG cohort.

| **Variables** | **SF wildtype** | **SRSF2** | **P** | **U2AF1** | **P** | **SF3B1** | **P** |
| --- | --- | --- | --- | --- | --- | --- | --- |
| No. of patients | 903 | 133 | - | 38 | - | 45 | - |
| Age, years, median (range) | 55 (18-86) | 65 (25-80) | **<0.001** | 64 (23-74) | **0.007** | 65 (31-78) | **0.001** |
| Female sex, no. (%) | 490 (54.3) | 31 (23.3) | **<0.001** | 9 (23.7) | **0.003** | 19 (42.2) | 0.387 |
| Hemoglobin, g/dL, median (range) | 9 (3.5-16) | 8.9 (3.8-14.7) | 0.466 | 9.2 (6-13.6) | 1.000 | 8.9 (6.8-13.4) | 1.000 |
| WBC count, 10<sup>9</sup>/L, median (range) | 22.4 (0.1-798.2) | 13.3 (0.5-406) | **0.008** | 7 (0.7-666) | 0.079 | 22.4 (0.9-269.5) | 1.000 |
| Platelets, 10<sup>9</sup>/L, median (range) | 55 (0-1760) | 49.5 (0-643) | 0.736 | 47 (11-132) | 0.466 | 67 (5-585) | 0.744 |
| LDH, U/L, median (range) | 448 (76-19624) | 362 (150-14332) | 0.118 | 346 (128-3085) | 0.313 | 472 (142-7434) | 0.950 |
| BM Blasts, %, median (range) | 80 (6-100) | 76 (15-100) | 0.455 | 60 (10-95) | **0.002** | 70 (13-95) | 0.206 |
| Performance Status (ECOG) > 1, no. (%) | 157 (25.9) | 18 (26.1) | 1.000 | 8 (27.6) | 1.000 | 4 (16) | 0.941 |
| primary AML, no. (%) | 786 (87) | 100 (75.2) | **0.016** | 24 (63.2) | **0.003** | 29 (64.4) | **0.002** |
| secondary AML, no (%) | 69 (7.6) | 30 (22.6) | **<0.001** | 13 (34.2) | **<0.001** | 11 (24.4) | **0.021** |
| therapy-related AML, no (%) | 48 (5.3) | 3 (2.3) | 0.507 | 1 (2.6) | 1.000 | 5 (11.1) | 0.380 |
| Allogeneic transplant, no. (%) | 296 (32.8) | 27 (20.3) | **0.022** | 7 (18.4) | 0.273 | 12 (26.7) | 0.797 |
| Complete Remission, no. (%) | 641 (71) | 71 (53.4) | **<0.001** | 18 (47.4) | **0.015** | 19 (42.2) | **0.001** |
| Relapse, no (%) | 366 (63.5) | 46 (82.1) | **0.034** | 11 (78.6) | 0.713 | 14 (87.5) | 0.266 |
| Deceased, no (%) | 575 (63.7) | 112 (84.2) | **<0.001** | 34 (89.5) | **0.010** | 41 (91.1) | **0.002** |

WBC: white blood cells, LDH: lactate dehydrogenase, BM: bone marrow, ECOG: Eastern Cooperative Oncology Group performance score

**Figure 1:**
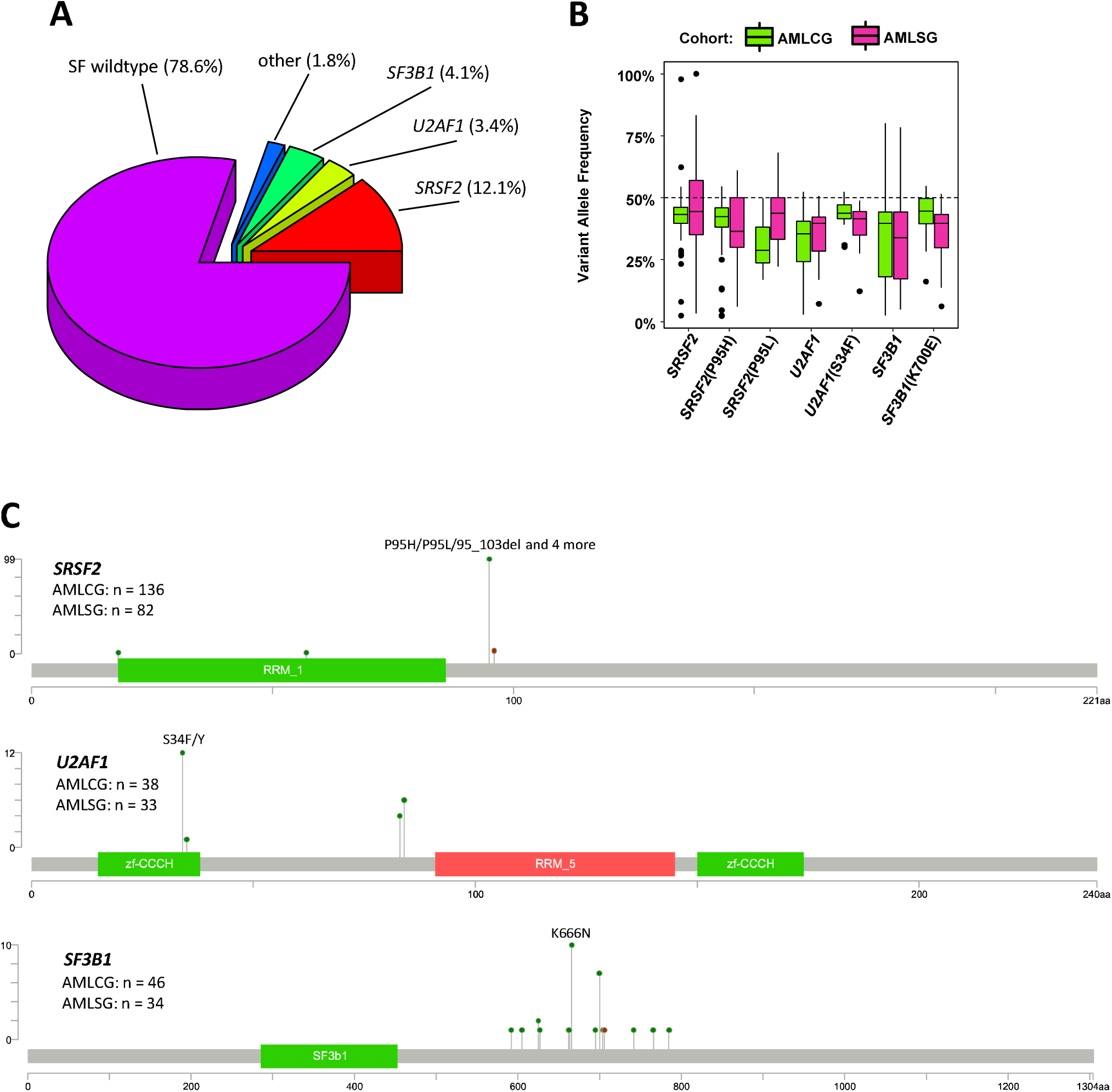
Frequency and location of SF mutations. (A) Distribution of SF mutations in the AMLCG cohort. (B) Variant allele frequency of SF mutations in both study cohorts. (C) Mutation plots showing the protein location of all SF mutations in the genes *SRSF2, U2AF1*, and *SF3B1* for all patients of the AMLCG cohort. Number of patients harboring SF mutations are additionally provided for both cohorts.

### Associations of SF mutations and other recurrent alterations in AML

In a second step, we investigated associations between SF mutations and recurrent cytogenetic abnormalities and gene mutations in AML (Figure 2). Notably, SF mutations were not found in inv(16)/t(16;16) patients (n=124), with the exception of one inv(16)/t(16;16) patient harboring a *U2AF1*(R35Q) mutation. The same held true for t(8;21) patients (n=98), where only one patient had a rare deletion in *SRSF2*. Additionally, all patients in the AMLCG cohort presenting with an isolated trisomy 13 (n=9) also harbored an *SRSF2* mutation (p<0.001), as described previously [31].

**Figure 2:**
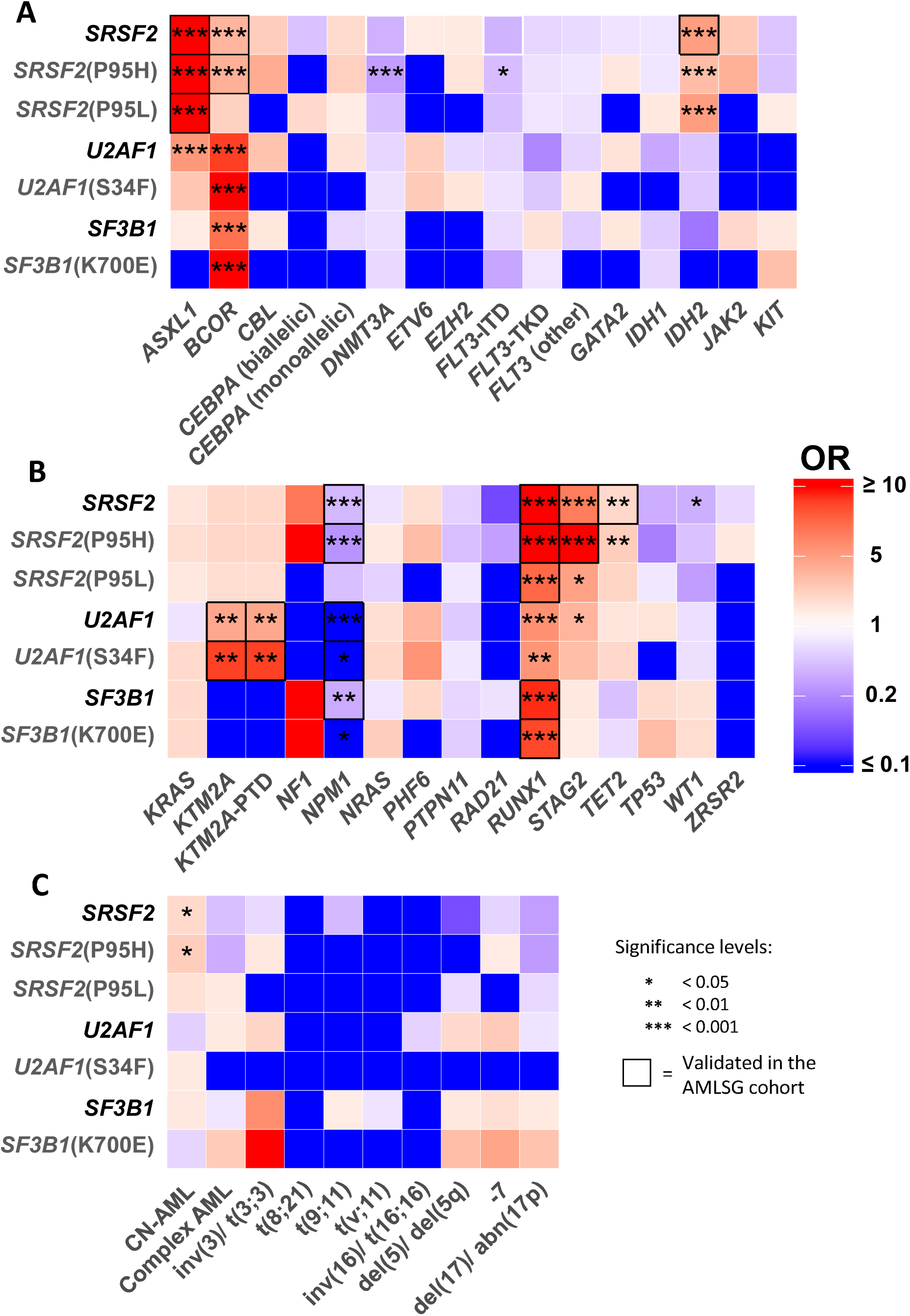
Correlations between SF mutations and recurrent abnormalities. Correlation matrix depicting the co-occurrence of SF mutations and recurrent mutations in AML (A), as well as cytogenetic groups, as defined in the ELN 2017 classification (B). Only variables with a frequency >1% in the AMLCG cohort are shown. FLT3-ITD: *FLT3* internal tandem duplication mutation; FLT3-TKD: *FLT3* tyrosine kinase domain mutation; CN-AML: cytogenetically normal AML

Mutations in all SF genes correlated positively with mutations in *BCOR* and *RUNX1* and negatively with mutations in *NPM1*. Expectedly, *SRSF2*(P95H) and *SRSF2*(P95L) mutations shared a similar pattern of co-expression including significant pairwise associations with mutations in *ASXL1, IDH2, RUNX1* (both p<0.001) and *STAG2* (p<0.001 and p=0.002, respectively). However, apart from *IDH2* mutations where co-occurrence was comparable (OR: 3.4 vs 5.1), mutations in *ASXL1, RUNX1* and *STAG2* coincided more frequently with *SRSF2*(P95H) mutations. Despite this, *SRSF2*(P95L) mutations showed a slightly increased co-occurrence with other recurrent AML mutations (median 5 vs 4 mutations, p=0.046).

### Prognostic relevance of SF mutations for relapse-free survival and overall survival

The prognostic impact of *SRSF2, U2AF1* and *SF3B1* mutations was initially assessed using Kaplan-Meier graphs and log-rank testing. All SF mutations presented with both inferior relapse-free survival (RFS) and overall survival (OS) compared to SF wildtype patients (Figures S3.1A-C and S3.2). The effect was most pronounced in *U2AF1* mutated patients with an one-year survival rate of only 29.1%, followed by *SF3B1* (40.6%) and *SRSF2* mutated patients (49.2%). Different point mutations inside the same SF gene did not differ significantly in their effect on OS.

To confirm the observed prognostic impact of SF mutations we performed single Cox regressions on all available clinical and genetic parameters. In agreement with the Kaplan-Meier estimates, patients harboring *SRSF2*(P95H), *SRSF2*(P95L), *U2AF1*(S34F) and *SF3B1*(K700E) mutations had significantly reduced RFS and OS (Figures S3.1D and S4.1). To test whether any SF mutation was an independent prognostic marker, multiple Cox regression models (Figure 3) were built by integrating all parameters significantly associated (p < 0.1) with RFS and OS in the single Cox regression models. Along with several known predictors, only *U2AF1*(S34F) mutations presented with prognostic relevance for both RFS (Hazard Ratio=2.81, p=0.012) and OS (HR=1.90, p=0.034) in the AMLCG cohort, but not in the AMLSG cohort. However, when aggregating mutations at the gene level, mutations in *SRSF2* and *SF3B1* presented with prognostic relevance for RFS in the AMLSG cohort (HR=1.77, p=0.008; HR=2.15, p=0.014; respectively), while not reaching significance in the AMLCG cohort (Table S4.2). When looking only at de novo AML patients, the prognostic impact of *U2AF1*(S34F) mutations diminished, yet the prognostic impact observed for *SRSF2* and *SF3B1* remained significant in the AMLSG cohort (HR=1.84, p=0.009; HR=2.43, p=0.015; respectively) (Tables S4.1-S4.2).

**Figure 3:**
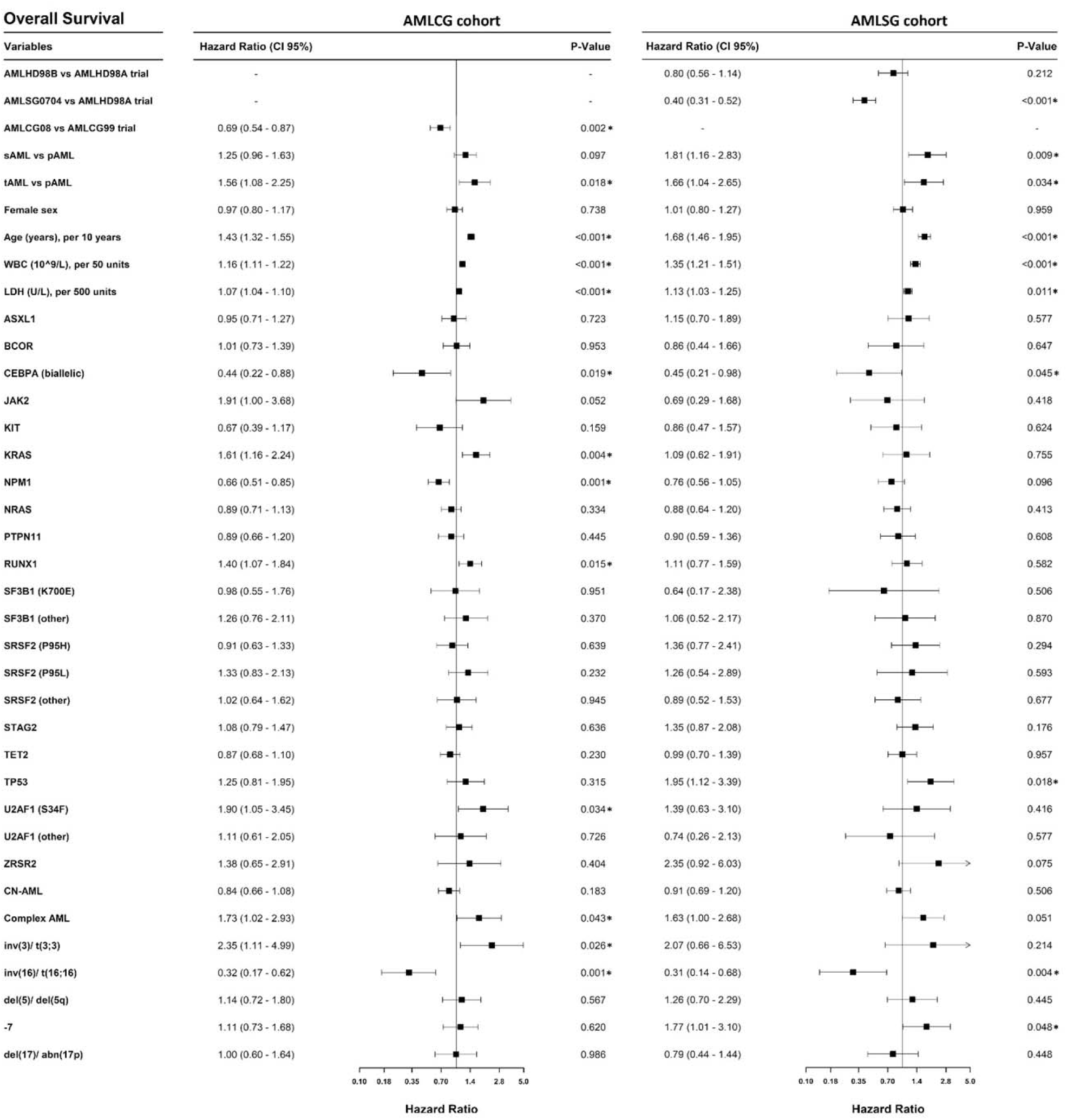
Multiple Cox regression models for overall survival. Multiple Cox regression models (for OS) were performed separately for patients in the AMLCG cohort (on the left) and AMLSG cohort (on the right). The models include all variables with *p* < 0.1 in the single Cox regression models of the primary cohort (AMLCG cohort). Significant p-values (<0.05) are marked with a star. CN-AML: cytogenetically normal AML; LDH: lactate dehydrogenase; pAML: primary AML; sAML: secondary AML; tAML: therapy-related AML; WBC: white blood cells

### Differential isoform expression in SF mutated patients

We next assessed the impact of SF mutations on mRNA expression. To this end, whole-transcriptome RNA-sequencing was performed on 246 AML patients, 29 of which harbored a mutation in the SF genes of interest, while 199 SF wildtype patients were used as a control (Figure S2). The remaining patients either presented with a different SF mutation (n=17) or exhibited more than one SF mutation (n=1) and were excluded. In addition, a subset of the Beat AML cohort (n=177) with matched DNA- and RNA-sequencing data was used for validation [20].

After low-coverage filtering we performed a differential isoform expression analysis for ∼90 000 isoforms. Differential expression was restricted to a small fraction of all expressed isoforms (<0.5%; Figure 4A and Table S6.1). Little overlap of differentially expressed (DE) isoforms was found when different SF mutation groups were compared to the control, consistent with previous observations [32]. However, 10 isoforms were reported as DE in both *SRSF2*(P95H) and *SRSF2*(P95L) mutated samples, all with the same fold-change direction (Figure 4B). Out of those, the isoforms in *GTF2I, H1F0, INHBC, LAMC1* and one of the isoforms of *METTL22* (*ENST00000562151*) were also significant in the validation cohort for both *SRSF2*(P95H) and *SRSF2*(P95L). Additionally, the isoform of *H1F0* was also reported as DE for *U2AF1*(S34F) mutants in both cohorts. For *SRSF2*(P95H) mutants 107 of all DE isoforms also reached significance in the validation cohort (40.1%), while for the other SF mutation subgroups validation rates ranged from 15.1 to 27.3% increasing with larger mutant sample sizes. Notably, mutated and wildtype samples showed large differences in the expression levels of several isoforms (Figure 4C and Figure S5). The top two overexpressed isoforms in *SRSF2*(P95H) both corresponded to *INTS3*, which was recently reported as dysregulated in *SRSF2*(P95H) mutants co-expressing *IDH2* mutations [33]. Several DE isoforms identified in SF mutated patients correspond to cancer-related genes, many of which have a known role in AML. Specifically, genes with DE isoforms included, but were not limited to *BRD4* [34], *EWSR1* [35] and *YBX1* [36] in *SRSF2*(P95H) mutated samples, *CUX1* [37], *DEK* [15,38] and *EZH1* [39] in *U2AF1*(S34F) mutated samples, as well as *PTK2* [40] in *SF3B1*(K700E) mutated patients (Tables S7.1-S7.2).

**Figure 4:**
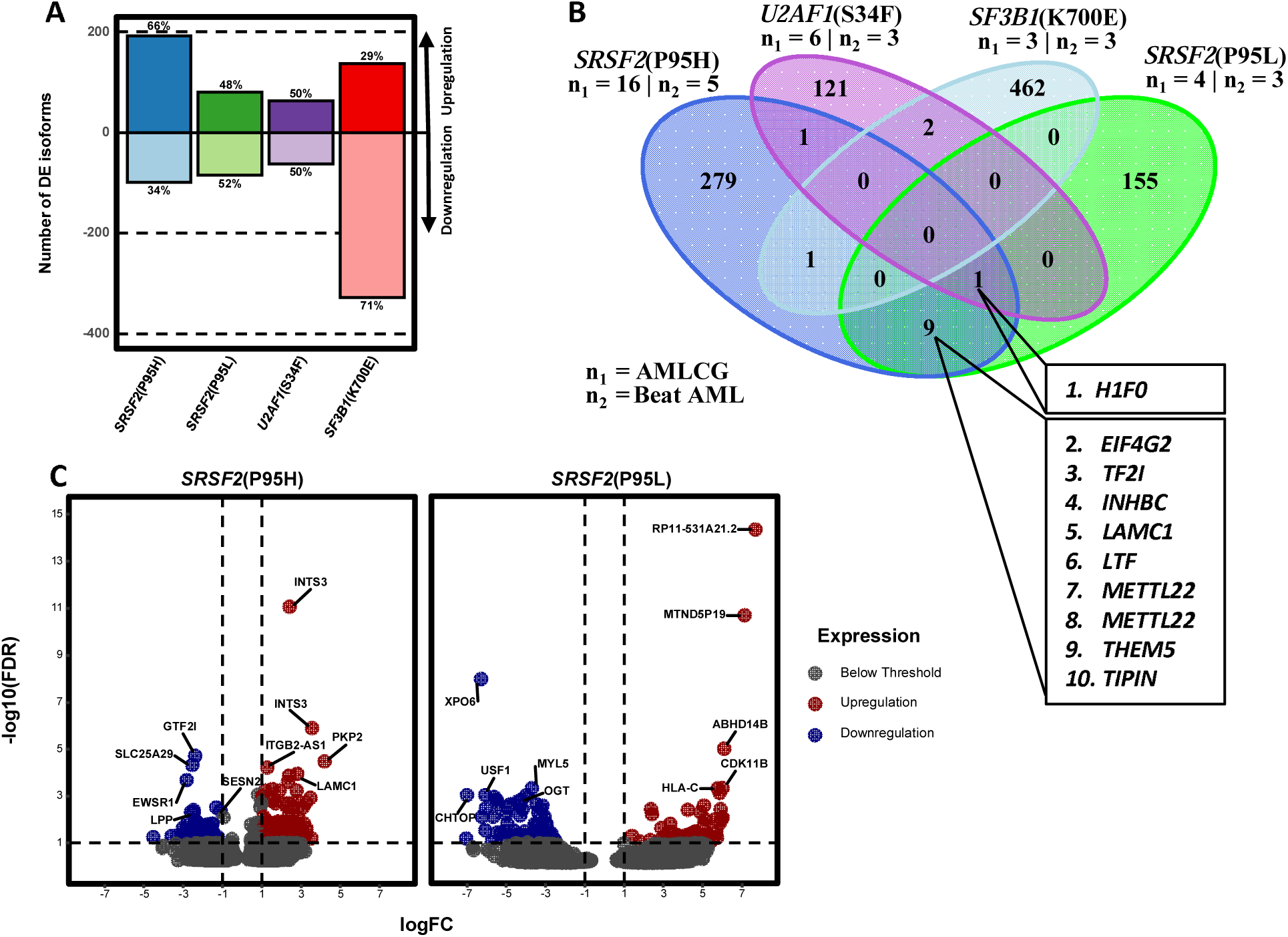
Differential isoform expression analysis in the AMLCG cohort. (A) Number of differentially expressed isoforms for each SF point mutation. (B) Isoforms reported as differentially expressed in the same direction among patients with different SF mutations vs. SF wildtype patients. HGNC symbols of the genes in which the common isoforms are located are shown. The corresponding Ensembl isoform identifiers are: 1. ENST00000340857, 2.ENST00000481621, 3. ENST00000309668, 4. ENST00000258341, 5. ENST00000426532, 6.ENST00000562151, 7. ENST00000564133, 8. ENST00000368817, 9. ENST00000566524, 10. ENST00000530211. (C) Volcano plots showing the magnitude of differential isoform expression for *SRSF2*(P95H) and *SRSF2*(P95L). The x-axis corresponds to the log_2_(fold change) of each isoform between mutated and wildtype samples, while the y-axis corresponds to –log10(FDR), where FDR represents the adjusted p-value for each isoform (False Discovery Rate).

Hierarchical clustering using DE isoforms was performed for all samples to assess the expression homogeneity of SF mutations. A tight clustering of samples harboring identical SF point mutations was observed, indicating an isoform expression profile highly characteristic for each individual SF mutation (Figures S6.1-S6.3). When using DE isoforms resulting from the comparison of all *SRSF2* mutated samples against SF wildtype samples, the samples did not cluster as well. This stands in agreement with the limited overlap of differentially expressed isoforms found between the two *SRSF2* point mutations examined and suggests at least some heterogeneity among them. The same also held true for *U2AF1* mutated samples, however all *SF3B1* mutated samples still clustered together when compared as a single group to the control.

### Differential splicing in SF mutants

Previous studies have reported differential splicing as causal for isoform dysregulation in SF mutants [41,42]. To detect aberrant splicing in our dataset, we quantified the usage of all unique splice junctions by pooling information from all samples (Figure S7). After filtering out junctions with low expression, 235 730 junctions were assigned to genes. Only junctions within an annotated gene were considered, leading to the exclusion of 11 617 (4.9%) junctions. Junctions present in genes with low-expression or genes with a single junction were excluded, leaving 221 249 unique junctions (19.3% novel) across 15 526 genes (Table S6.1). Applying the same workflow to the Beat AML cohort yielded 194 158 junctions (8.3% novel). Notably, of the 172 518 junctions shared across both datasets, 10 029 (5.8%) were novel. The novel junctions passing our filtering criteria were supported by a high amount of reads and samples with a distribution comparable to that of annotated junctions (Figure 5A). Neither the number of novel junctions nor the number of reads supporting them correlated with the presence of SF mutations, suggesting that novel splicing events are not increased in SF mutants.

**Figure 5:**
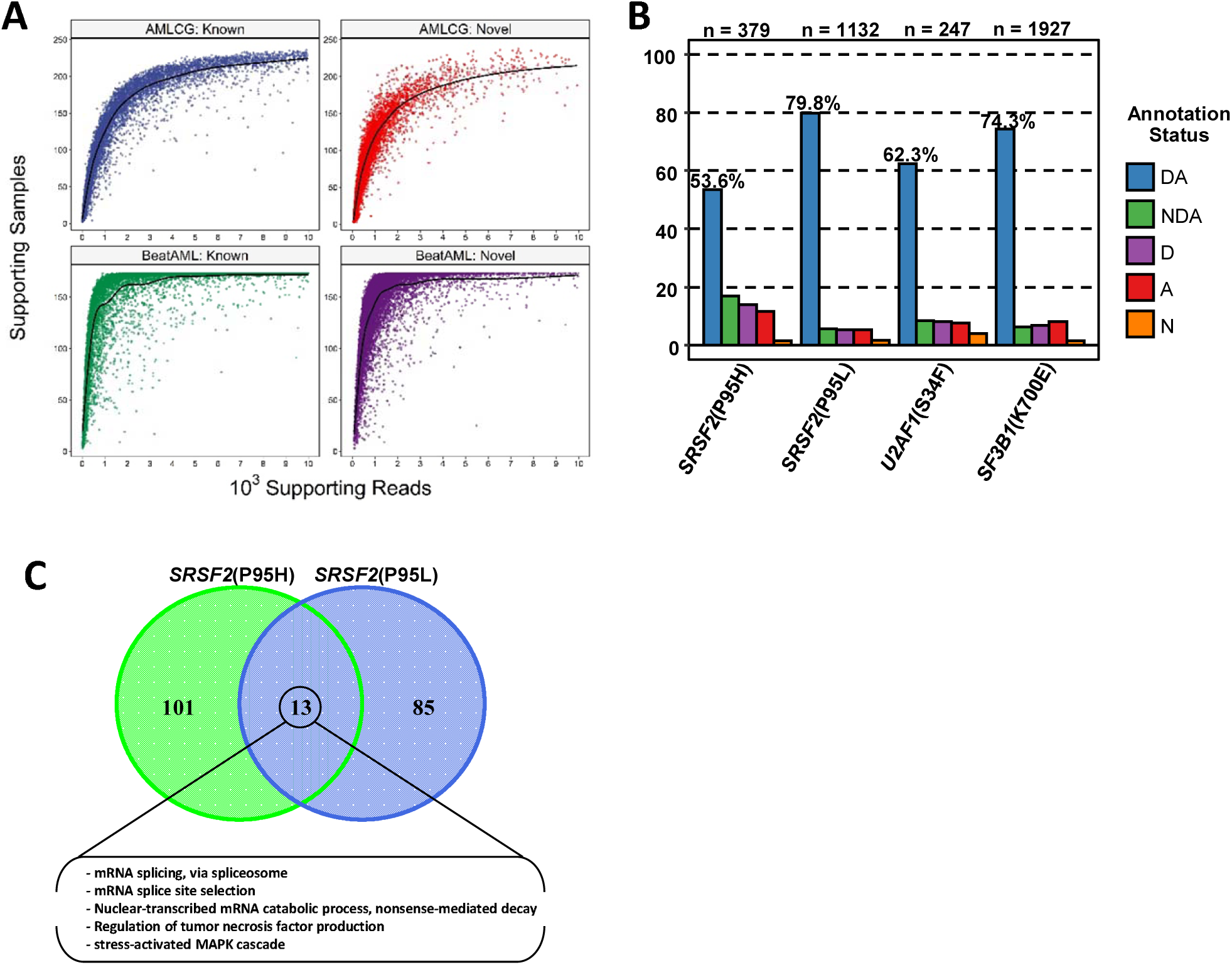
Differential splice junction usage. (A) Scatterplot displaying the number of samples as well as the total number of reads supporting each splice junction, separately for known and novel splice junctions in both RNA-Seq datasets. To preserve visibility 20000 random junctions are shown for each group. (B) Barchart showing the annotation status of splice junctions reported as differentially used. Novel splice junctions where classified into 5 groups based on their annotation status as described previously[15] (“DA”: annotated junctions, “NDA”: unknown combination of known donor and acceptor sites, “D”: known donor, but novel acceptor site, “A”: novel donor, but known acceptor site, “N”: previously unknown donor and acceptor site). (C) Venn diagram showing the overlap of GO terms between *SRSF2*(P95H) and *SRSF2*(P95L) mutants for the “biogical process” domain.

In consideration of the high proportion of novel junctions in both datasets, we employed a customized pipeline that can quantify the differential splice junction usage (DSJU) of each individual junction, by harnessing usage information from all junctions inside one gene. Of the several hundred junctions reported as differentially used in our primary cohort (p<0.05, log_2_(fold change) > 1), 20.2-45.9% constituted novel junctions (Tables S7.1-S7.3 and Tables S9.1-S9.2) and were classified according to their relationship with known acceptor and donor sites as described previously (Figure 5B) [15]. Unsurprisingly, validation rates increased with larger mutant sample sizes, ranging from 9.3% (*SF3B1*(K700E); n = 3) to 74.0% (all *SRSF2* mutants; n = 26). Furthermore, validation rates were higher for novel junctions (mean 39.3% vs. 21.5% known junctions), likely due to the stricter initial filtering criteria applied. By performing nanopore sequencing of one *SRSF2*(P95H) mutant and one SF wildtype sample we were able to confirm the usage of several novel junctions and detect resulting novel isoforms as exemplified for *IDH3G* in Figure 6A-D. A tendency towards decreased junction usage was observed for all SF point mutations and was most evident in *SF3B1*(K700E) mutants (1 423/1 927; 73.9% of differentially used junctions). The total number of splicing events, however, was not reduced in SF mutants (mean 9,275,359 events vs. 9 192 697 in wildtype patients), suggesting that decreased splicing is limited to selected junctions rather than being a global effect.

**Figure 6:**
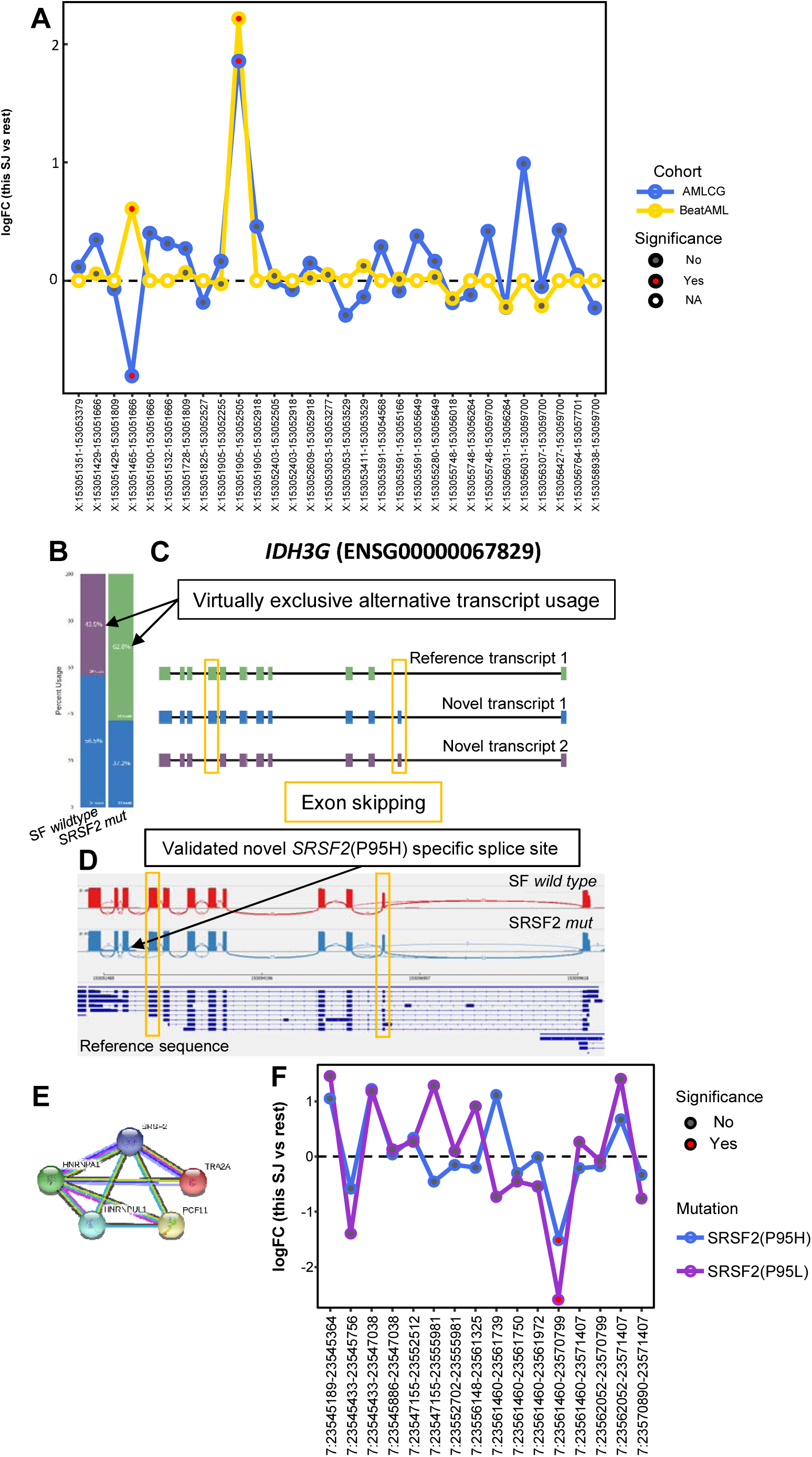
Splicing dysregulation in *SRSF2* mutants. (A) DSJU of all splice junctions inside *IDH3G* (ENSG00000067829) are shown for *SRSF2*(P95H) mutants compared to SF wildtype patients in the AMLCG and Beat AML cohort. To determine significance the log fold-change (logFC) of each splice junction (SJ) is compared to the logFC of all other junctions inside the same gene. The x-axis denotes individual splice junctions defined by their chromosomal coordinates (note the high number of splice sites shared by multiple splice junctions). (B)-(D) Nanopore sequencing results of one *SRSF2*(P95H) mutated sample and one SF wildtype sample of the AMLCG cohort. The yellow boxes highlight examples of exon skipping (same exons highlighted in (C) and (D). (B) FLAIR distribution of transcripts. Only one known isoform is expressed in the samples. Additionally, two novel isoforms were detected which are virtually mutually exclusive in the *SRSF2*(P95H) mutant and the SF wildtype sample. (C) Exon composition of known and novel isoforms detected (D). Sashimi plots showing the exon sequence coverage as well as the splice junction usage of the *SRSF2*(P95H) mutated and SF wildtype samples. The black arrow indicates the novel splice junction that is differentially used in *SRSF2*(P95H) mutated samples compared to SF wildtype samples in both RNA-Seq datasets shown in (A). (E) STRING plot depicting an interaction between SRSF2 and several other splicing-related proteins including *TRA2A*. (F) Differential splice junction usage of all splice junctions inside *TRA2A* (ENSG00000164548) are shown for *SRSF2*(P95H) and *SRSF2*(P95L) mutants compared to SF wildtype patients. Annotation same as (A).

We systematically compared the genes with at least one DE isoform and those reported as differentially spliced in all SF mutation subgroups (Tables S9.3-S9.4). For *SRSF2* mutants, genes significant in both analyses included *EWSR1, H1F0, INTS3* and *YBX1*. In general, out of the genes examined in both analyses only 9.8-23.3% (depending on the SF mutation) of genes reported as having a DE isoform were also reported as being differentially spliced. Conversely, 3.3-28.5% of differentially spliced genes were also reported as having a DE isoform. These findings suggest that differential gene splicing does not always lead to altered isoform expression while at the same time differential isoform expression cannot always be attributed to an explicit splicing alteration. Considering the complementary nature of the analyses, we performed gene ontology (GO) analysis by combining the genes with evidence of differential isoform usage or differential splicing (Tables S10.1-S10.7). Interestingly, GO terms enriched for both *SRSF2* mutants included “mRNA splicing, via spliceosome” (p<0.001 and p=0.046, respectively) and “mRNA splice site selection” (p=0.022 and p=0.019, respectively) (Figure 5C).

In an additional step, the splice junction counts reported by *Okeyo-Owuor et al*. were used to detect DSJU between CD34+ cells with U2AF1(S34) mutations (n=3) and SF wildtype (n=3) via the same pipeline applied to the AMLCG and Beat AML cohorts. While no identical junctions were differentially used in all three datasets, 16 genes were reported as differentially spliced in all, including leukemia or cancer-associated genes (*ABI1, DEK, HP1BP3, MCM3* and *SET*), as well as *HNRNPK* (a major pre-mRNA binding protein), thereby further refining our list of genes with strong evidence of differential splicing between *U2AF1*(S34F) mutants and SF wildtype samples (Table S11).

### The effect of SF mutations on the splicing pathway

It is well established that SF mutations dysregulate splicing by changing RNA binding affinities or altering 3’ splice site recognition of the corresponding splicing factors. However, SRSF2 also plays a splicing-independent role in transcriptional pausing by translocating the positive transcription elongation factor complex (P-TEFb) from the 7SK complex to RNA polymerase II [43]. A recent study reported that mutant SRSF2(P95H) enhances R-loop formation due to impaired transcriptional pause release, thus providing evidence that altered splicing does not account for the entirety of the *SRSF2*(P95H) mutant phenotype [32]. In another study no difference was found in the total mRNA of 12 Serine/arginine rich splicing factors and 14 out of 16 major heterogeneous nuclear ribonucleoprotein (hnRNP) splicing factors between *SRSF2*(P95H) mutant and WT CRISPR clones [44].

We cross-referenced our differential expression and differential splicing analysis results with a list of all genes involved in splicing (GO:0000398; mRNA splicing, via spliceosome). Of the 347 splicing-related genes (317 of which were expressed in our dataset) 101 were dysregulated in at least one SF mutant group. On average 30.5 (range 6-52) splicing-related genes were dysregulated per SF point mutation. Of note, both *SRSF2* point mutations associated with differential splicing of *HNRNPA1* and *HNRNPUL1*, as well as *PCF11* and *TRA2A*. A query of the STRING database suggests protein-protein interactions between SRSF2 and the proteins of the above genes, which are also interconnected (Figure 6E). Interestingly, one of the differential splicing events reported in both *SRSF2* mutants involves the under-usage of the same novel splice junction in *TRA2A* (Figure 6F). *TRA2A* has previously been shown to be differentially spliced in mouse embryo fibroblasts upon *SRSF2* knockout [45]. Furthermore, it has been shown that both HNRNPA1 and SRSF2 interact with the loop 3 region of 7SK RNA and by favoring the dissociation of SRSF2, HNRNPA1 may lead to the release of active P-TEFb [46]. Taken together, our results indicate a strong dysregulation of the splicing pathway in SF mutants including several genes whose gene products closely interact with SRSF2.

## Discussion

The clinical relevance of SF mutations and their aberrant splicing patterns have been explored in myelodysplasia, while comparable data for AML is lacking. In this study we examined two AML patient cohorts, encompassing a total of 2678 patients from randomized prospective trials, to characterize SF mutations clinically. This analysis was complemented by RNA-sequencing analysis of two large datasets to reveal targets of aberrant splicing in AML. We show that SF mutations are frequent alterations in AML, identified in 21.4% of our primary patient cohort, especially in elderly patients and in secondary AML. SF mutations are associated with other recurrent mutations in AML, such as *BCOR* and *RUNX1* mutations, however *SRSF2*(P95L) mutations co-occur less often with those mutations when compared to *SRSF2*(P95H) mutations, albeit showing a slightly increased mutational load. This suggests a more diverse co-expression profile of *SRSF2*(P95L).

Previous studies have demonstrated the predictive value of SF mutations in clonal haematopoiesis of indeterminate potential (CHIP) [47], MDS [6,8,48–50] and AML [10,18,19,51]. However, survival analyses in AML were, in their majority, hampered by small sample sizes and limited availability of further risk factors. Therefore, we examined whether SF mutations impact survival while accounting for recently proposed risk parameters included in the ELN 2017 classification [52]. In our analysis, *SRSF2* and *SF3B1* mutations were no independent prognostic markers for OS in AML. *U2AF1*(S34F) mutations displayed poor OS in the AMLCG cohort, which we were unable to validate in the AMLSG cohort. The discrepancy in survival of SF mutated patients between the two cohorts lied most likely in the large age difference of the participants (median age difference of 8 years), which also led to a higher percentage of patients receiving allogeneic transplants in the AMLSG cohort (56.5% vs. 30.6% in the AMLCG cohort). In summary, SF mutations are early evolutionary events and define prognosis and transformation risk in CHIP and MDS patients, yet there is no clear independent prognostic value of SF mutations in AML.

Two large RNA-sequencing studies have been performed previously, to detect aberrantly spliced genes in SF mutants, both of which focused on MDS patients [41,42]. In this study we described a distinct differential isoform expression profile for each SF point mutation. Furthermore, we evaluated differential splicing for the four most common SF point mutations via a customized pipeline to determine differential usage of both known and novel splice junctions. Our pipeline enables the differential quantification of individual splice junctions without restricting the analysis to annotated alternative splicing events. We argue that the strength of our analysis lies in the accurate detection of single dysregulated junctions (especially in cases where splice sites are shared by multiple junctions) in an annotation-independent manner achieving validation rates up to 74.0% in our largest mutant sample group (*SRSF2*, n=19). Limitations of the analysis include the restriction to junctions with both splice sites within the same gene (a restriction shared by most differential splicing algorithms) and genes with at least two junctions. However, the reduced requirements of our analysis could prove valuable in the study of differential splicing in organisms with lacking annotation. All SF point mutations shared a tendency towards decreased splice junction usage, which did not affect the global number of splicing events in SF mutants. Surprisingly, we observed a limited overlap between genes with differentially expressed isoforms and differentially spliced genes. In addition, a recent study by *Liang et al*. reported that the majority of differential binding events in *SRSF2*(P95H) mutants do not translate to alternative splicing [53]. Taken together, these findings indicate a “selection” or possibly a compensation of deregulatory events from differential binding through differential splicing to finally differential isoform expression. Furthermore, the enrichment of aberrant splicing in splicing-related genes opens the possibility of a cascading effect on transcription via the differential alternative splicing of transcriptional components. A congruent hypothesis was stated by *Liang et al*., where an enrichment of *SRSF2*(P95H) targets in RNA processing and splicing was shown, further supporting the notion of an indirect effect of mutant SRSF2 facilitated through additional splicing components. Further investigations may provide a mechanistic link between the differential splicing of selected genes and the impairment of transcription and specifically transcriptional pausing observed in SF mutant cells, which contributes to the MDS phenotype [32].

To the best of our knowledge our study represents the most comprehensive analysis of SF mutations in AML to date, both in terms of clinical and functional characterization. This enabled us to study *SRSF2*(P95H) and *SRSF2*(P95L) separately, thereby not only outlining their differences but also identifying common and likely core targets of differential splicing in *SRSF2* mutants. We conclude that SF mutated patients represent a distinct subgroup of AML patients with poor prognosis that is not attributable solely to the presence of SF mutations. SF mutations induce aberrant splicing throughout the genome including the dysregulation of several genes associated with AML pathogenesis, as well as a number of genes with immediate, functional implications on splicing and transcription. Further studies are required to identify which splicing events are critical in leukaemogenesis and whether they are accessible to new treatments options, such as splicing inhibitors [54] and immunotherapeutic approaches.

## Data Availability

The raw data of this study is available, but German law restrictions apply to the availability of these data, which were used for the current study, and so are not publicly available. Data are however available from the authors upon reasonable request and with permission of the local ethics committee.

https://data.mendeley.com/datasets/7sg6bn8m74/draft?a=57983ef4-798a-4ade-ac94-8304628cf27a

## Acknowledgments

The authors thank all participants and recruiting centers of the AMLCG, BEAT and AMLSG trials.

## Author Contributions

S.A.B., A.M.N.B. and T.H. conceived and designed the analysis. S.A.B., A.M.N.B., V.J., M.R.-T., H.J., A.G., S.C., N.K., K.S., K.H.M. and T.H. provided and analyzed data. A.M.N.B., V.J. and U.M. provided bioinformatics support. J.P.-M., S.K. and H.B. managed the Genome Analyzer IIx platform and the RNA-sequencing of the AMLCG samples. M.R.-T., H.J., B.K., S.S., N.K., S.K.B., K.H.M. and K.S. characterized patient samples; M.C.S., D.G., W.B., B.W.,J.B. and W.H. coordinated the AMLCG clinical trials. S.A.B. and T.H. wrote the manuscript. All authors approved the final manuscript.

## Additional Information

The authors declare no conflicts of interest.

